# GWAS links *APOE* to neuropsychiatric symptoms in mild cognitive impairment and dementia

**DOI:** 10.1101/2025.01.31.25321498

**Authors:** Selina M. Vattathil, Freida Blostein, Tyne W. Miller-Fleming, Lea K. Davis, Alzheimer’s Disease Genetics Consortium (ADGC), Alzheimer’s Disease Neuroimaging Initiative, Thomas S. Wingo, Aliza P. Wingo

## Abstract

**INTRODUCTION:** Neuropsychiatric symptoms in dementia (NPS) collectively refer to behavioral and psychological symptoms affecting individuals with mild cognitive impairment (MCI) or Alzheimer’s disease or related dementia (ADRD). NPS are among the most troubling aspects of living with dementia and their treatments have limited efficacy. We aim to investigate genetic variants contributing to NPS to identify new therapeutic targets.

**METHODS:** We performed a genome-wide association study (GWAS) for nine NPS domains measured by the NPI-Q in 12,800 participants of European ancestry with MCI or ADRD recruited by Alzheimer’s disease research centers across the U.S.

**RESULTS:** We found genome-wide significant signals for agitation, anxiety, apathy, delusions, and hallucinations in the *APOE* locus that were driven by the *APOE* ε4 allele. We replicated these findings in two independent datasets. Mediation analyses revealed that MCI/ADRD severity only partially mediated the GWAS signals, except for apathy.

**DISCUSSION:** These findings suggest the *APOE* ε4 allele influences NPS independently of and beyond its effect on ADRD.

## 1 Introduction

Neuropsychiatric symptoms in dementia (NPS) encompass a range of behavioral and psychological symptoms affecting approximately 60% of individuals with mild cognitive impairment (MCI) and 75% of those with dementia^1–3^. These symptoms, sometimes referred to as behavioral and psychological symptoms of dementia, include apathy, depression, anxiety, irritability, agitation, sleep disturbance, delusions, and hallucinations^1,2^ and are associated with greater functional impairment, higher caregiver burden, and earlier institutionalization^4–6^. Despite their prevalence and impact, current therapeutics have limited efficacy^4,7,8^ and can cause substantial side effects^9,10^, underscoring the urgent need for safe and effective treatments.

In dementia, cognitive performance typically shows a persistent and gradual decline, while NPS often manifest episodically and vary with dementia severity—affective symptoms are more common in early stages while delusions and hallucinations emerge in later stages. Nevertheless, NPS are associated with faster cognitive decline^4,11–14^. Understanding the extent to which NPS and cognitive impairment stem from shared or distinct pathophysiology is critical for developing targeted and effective treatments for the various symptoms experienced by each patient.

Studying the genetic basis of NPS can illuminate disease mechanisms and potential therapeutic targets, but genetic studies of NPS are currently limited. The overlap between NPS and psychiatric conditions like bipolar disorder and major depressive disorder—both of which share genetic heritability with Alzheimer’s dementia (AD)^15^—has prompted candidate gene studies exploring genes linked to neurodevelopment, neurotransmitter systems, and AD^16–20^. Among these, *APOE* stands out as the strongest candidate gene due to its well-established association with AD, but evidence for its involvement in NPS has been mixed^21,22^. The studies generally had small sample sizes, leading to low statistical power and false positives. Candidate gene studies have lost precedence in recent years to agnostic genome-wide association studies (GWAS), and two GWAS for NPS have been conducted to date. These GWAS investigated specific NPS subdomains, namely psychosis (hallucinations or delusions) and affective symptoms (anxiety, depression, or irritability), and identified three risk loci (*ENPP6*, *SUMF1*, and *APOE*) for psychosis^23–25^.

To further investigate genetic variants contributing to NPS, we conducted GWAS for nine individual NPS symptoms in 12,806 participants with MCI or dementia. This study differs from previous GWAS in two key features. First, the previous studies focused on psychosis and affective symptoms, which are aggregate phenotypes defined from combinations of individual NPS symptoms. We considered a wide range of NPS symptoms individually, which we hypothesized could reveal new loci since each symptom domain may arise through unique biology. Second, the previous studies were specific to participants with AD. We expanded the analyses to also include participants with MCI, since NPS and mild behavioral impairment can emerge in early cognitive impairment or even before cognitive decline^26^. We identified genome-wide significant associations between variants in *APOE* and several NPS, including agitation, anxiety, apathy, delusions, and hallucinations. While *APOE* is a known risk factor for cognitive decline, mediation analyses showed that it may also contribute directly to NPS.

## 2 Methods

### 2.1 Discovery Dataset

The discovery dataset were participants recruited from 32 Alzheimer’s Disease Research Centers (ADRCs) whose data was curated and merged into the Uniform Data Set (UDS)^27–30^ by the National Alzheimer’s Coordinating Center (NACC). Each participant underwent a comprehensive annual evaluation using established research diagnostic criteria to assign diagnosis^27–31^. For this study, de-identified data were obtained from NACC using the following inclusion criteria: diagnosis of MCI^32^ or dementia^33^ with complete data for age at visit and sex, available whole genome genotyping, and European genetic ancestry. Severity of cognitive impairment was treated as either a continuous variable or a binary variable, as noted for each analysis. As a continuous variable, severity was defined as the average global Clinical Dementia Rating (CDR) score across visits at which the participant experienced their most severe observations for the NPS domain. As a binary variable, severity was defined as either “dementia” or “MCI”.

### 2.2 Neuropsychiatric symptoms of dementia (NPS)

The discovery datasets have NPS domains measured longitudinally with the Neuropsychiatric Symptom Inventory Questionnaire (NPI-Q)^34^. Nine domains were considered in this study: agitation or aggression, anxiety, apathy or indifference, delusions, depression or dysphoria, disinhibition, hallucinations, irritability or lability, and nighttime behaviors. Annual assessments of NPS indicate presence and severity (e.g., mild, moderate, severe) in the preceding month.

For the GWAS analyses, we classified a participant as a ‘case’ if they experienced the symptom in the month preceding any visit, and as a ‘control’ otherwise. For characterizing the correlation between NPS and other traits, a semiquantitative score was defined for each participant based on their maximum observed NPS severity (none, mild, moderate, severe).

### 2.3 Genotyping and imputation

Genotyping, initial QC, and imputation were performed by ADGC. Initial QC included outlier removal and inspection of population substructure to obtain sets of relatively genetically similar participants. Specifically, genetic principal components (PCs) were computed for study participants and plotted against 1000 Genomes Phase 3 (1KG) reference samples. Samples not visually clustering with expected reference populations based on self-reported race and ethnicity were filtered. NACC participants reported race from the following choices: American Indian or Alaska Native, Asian, Native Hawaiian or Other Pacific Islander, Black or African American, White, Other, or Unknown. Participants also reported their ethnicity as Hispanic or Non-Hispanic. ADGC divided participants into four groups based on self-reported race and ethnicity: African-American, Asian, Hispanic, and non-Hispanic white. For participants who self-identified as non-Hispanic White, the expected 1KG reference populations were CEU and TSI. After PC-based filtering, imputation was performed using the TopMed server.

### 2.4 GWAS data preparation

Due to the small sample size in each of the African-American, Asian, and Hispanic datasets (< 1500 participants with MCI/dementia), our analysis focused on the non-Hispanic white dataset. We performed further genetic quality control by filtering the imputed genotype data to retain biallelic single-nucleotide polymorphisms (SNPs) with imputation r^2^ > 0.8, minor allele frequency > 1%, genotype missingness < 5%, and HWE p-value > 10^-8^, then removed any SNPs missing from one or more batches, leaving 7,708,866 autosomal and pseudoautosomal region (PAR) SNPs. Next, we assessed relatedness using KING and filtered out 162 individuals to yield 12,839 unrelated participants (no 2^nd^ degree or closer relatives). We then visualized batch effects and identified sample outliers using smartPCA (Eigensoft) and filtered 33 outliers, resulting in a final dataset of 12,806 participants.

Genetic ancestry was confirmed by projecting study samples onto the 1KG PC space using PLINK; all samples clustered with European ancestry populations, as expected based on the prior ADGC PC-based filtering.

### 2.5 GWAS analysis

We performed parallel logistic regressions using PLINK for each NPS domain while controlling for sex (male v. female), batch (categorical with 15 levels), and the first 10 genetic PCs. Keeping with the conventional significance threshold for GWAS of common variants, significant association was defined as p-value < 5×10^-8^.

### 2.6 Mediation analysis

Mediation analysis was performed in R using the package mediation^35^ with SNP as the treatment variable and cognitive impairment severity or age of onset of cognitive impairment as the mediator. Both the outcome model and mediator model were adjusted for sex, batch, and ten genetic PCs. P-values and confidence intervals were estimated using 10,000 bootstrapped simulations. Significant mediation was defined as average causal mediation effects (ACME) p-value < 0.05. In case of significant mediation, it was defined as partial mediation if the 95% confidence interval for the proportion mediated did not include 1 and a significant direct effect remained (p-value < 0.05), and complete mediation otherwise.

### 2.7 Replication in BioVU

#### 2.7.1 Identification of BioVU participants with mild cognitive impairment or dementia

We conducted a replication analysis in the Vanderbilt University Medical Center’s (VUMC) genetic biobank, called BioVU, and the accompanying deidentified version of the VUMC electronic health record, called the Synthetic Derivative (data freeze June 2023). The study was approved by VUMC IRB (IRB 201609). For this analysis we considered only participants with genotype data measured on the Illumina Expanded Multi-Ethnic Genotyping Array (MEGAex)^36^. Participants with genotype data had an average length of medical record of approximately 10 years. Since the discovery analysis was conducted among individuals of European ancestry, we restricted the replication analysis to individuals of European ancestry, where ancestral clusters for BioVU participants were defined as previously described using Eigenstrat and genotype data imputed from the MEGAex array and the Haplotype Reference Consortium reference panel using the Michigan Imputation Server^37,38^. Genotyping data was subjected to a series of ancestry-specific QC filters, including minor allele frequency<0.005, imputation quality *R*^2^ < 0.3 thresholding and 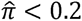. The resulting data set contained 6,360,678 variants from 66,917 participants. The data set was further restricted to individuals with non-missing demographic data on electronic health record reported sex and date of birth (n=66,457).

To define a cohort of individuals with MCI or dementia within this BioVU European ancestry population (“MCI/dementia cohort”), we selected any individual with at least one ICD9 or ICD10 code relating to dementia, mild cognitive impairment, or Alzheimer’s disease (**Supplementary Table 1**). We excluded individuals with a first recorded dementia or mild cognitive impairment ICD code at or before the age of 50, resulting in a final sample size of n=2456.

To verify our definition of MCI/dementia, we matched individuals in the MCI/dementia cohort to individuals in the overall European ancestry BioVU MEGAex cohort on median age across record and sex using propensity score matching and a 1:4 matching ratio. We then compared the distributions of alleles at rs429358 and rs7412, which determine *APOE* isoforms^39^.

Principal components were calculated among the MCI/dementia cohort on a randomly selected subset of 250,000 SNPs using flash PCA and an in-house script^40^.

#### 2.7.2 NPS outcomes

Individuals with dementia or MCI in the VUMC health system are not routinely assessed for the NPI-Q. Instead, we created a variable which roughly maps to underlying NPS constructs of the NPI-Q using the data types available in BioVU, namely, ICD codes and psychotropic medications.

We created a binary variable “any NPS” based on medication data in the medical record, specifically for receipt of antipsychotic or antidepressant medications (see **Supplementary Table 2** for list of medications). To identify individuals receiving these medications in response to neuropsychiatric symptoms stemming from dementia and/or MCI (i.e., “any NPS” case), we defined individuals with first recorded receipt of these medications within 365 days preceding or any time after their first diagnosis of MCI/dementia as a case; all other individuals were defined as not having NPS, or controls.

#### 2.7.3 Replication of GWAS findings

We performed replication analysis using the NPS variable described above for the two independent loci (based on LD threshold) with significant association signals in the discovery analysis. We extracted the SNPs with p-value < 5×10^-8^ from the discovery GWAS, resulting in 25 SNPs for testing. We performed logistic regression while controlling for sex and the first ten principal components of the genotype data using PLINK. We applied a Bonferroni correction for multiple testing, setting the significance threshold at 0.05/ [2 independent loci] = 0.025.

The BioVU genetic data was controlled for batch effects by removing any SNPs that have association with batch; however, as a sensitivity analysis we also controlled for batch. Because this analysis only included individuals with MCI/dementia by design, some batches had low cell counts in the sensitivity analysis and were dropped.

#### 2.7.4 Mediation analysis

We replicated the mediation analysis conducted in the discovery analysis. Specifically, we tested whether the association between the SNPs and NPS was mediated by cognitive impairment. We examined severity of cognitive impairments in two different ways: (i) dementia versus MCI diagnosis based on ICD codes and (ii) age of onset of dementia and/or MCI. For dementia versus MCI diagnosis, we defined individuals as having MCI if they only ever had the ICD code relating to mild cognitive impairment, i.e., G31.84, otherwise individuals were classified as having dementia. For age of onset, we used the age at the first recorded MCI/dementia ICD code. We then performed mediation analyses adjusting for sex and the first ten genetic principal components using the R package mediation^35^ with 1000 bootstrapped simulations.

### 2.8 Replication in ADNI

#### 2.8.1 ADNI cohort

We obtained data from the Alzheimer’s Disease Neuroimaging Initiative (ADNI) database (adni.loni.usc.edu). ADNI is a multisite program aimed at validating Alzheimer’s disease biomarkers. ADNI is a longitudinal study that enrolls participants aged 55 to 90 years who undergo standardized diagnostic assessments every six months. Assessments included clinical diagnosis of normal cognition, MCI, or dementia as well as NPS assessment using NPI and/or NPI-Q. A total of 1,566 samples underwent WGS, including some samples that were sequenced as part of a collaboration between ADNI and the ADSP Follow-up Study.

#### 2.8.2 Definition of cognitive impairment and NPS outcomes

For our replication analysis, we subsetted the data to the 1,059 participants who were diagnosed with MCI or dementia during the follow-up period and did not revert to normal cognition. For missing values in the longitudinal data on diagnosis of cognitive impairment and global CDR score, we imputed the missing value using the previous visit’s observation if it matched the observation from the subsequent visit. NPS case status was defined using NPI or NPI-Q responses following the same criteria as the discovery analysis, i.e., a participant was classified as a case if they experienced the symptom in the month preceding any visit, and as a control otherwise. Sex (male/female) was self-reported.

#### 2.8.3 WGS genotyping and quality control

Whole genome sequencing was performed using a HiSeq2000. Reads were mapped and variants were called against GRCh38. We restricted the data to biallelic SNPs, then removed variants with missingness > 10% and Hardy-Weinberg p-value < 10^-8^. Additionally, we removed 179 samples that were already analyzed as part of the discovery GWAS and two samples closer than third degree relatives. To match the discovery sample, we selected individuals of European genetic ancestry using the probabilistic principal components model-based inference tool from Bystro^41^, which predicts the most likely continent-level ancestry for each participant using reference data from 1000 Genomes populations. Eighty participants were removed in this step, leaving 798 participants for analysis.

#### 2.8.4 Replication of GWAS findings

We performed replication analysis for the six significant association signals from the discovery analysis, which included the association of the *APOE* locus with five NPS symptoms and the *ADAMTSL1* locus with one NPS symptom. We extracted the SNPs with p-value < 5×10^-8^ in the discovery analysis, resulting in 25 SNPs for testing. SNP association was tested using logistic regression controlling for sex and 10 genetic PCs using PLINK. We applied a Bonferroni correction for multiple testing, setting the significance threshold at 0.05 / [6 tests] = 8.33 × 10^-3^.

#### 2.8.5 Mediation analysis

Mediation analysis to test whether the SNP association with NPS was mediated by cognitive impairment severity or age of onset was performed with R package mediation^35^ using the same parameterizations as in the discovery analysis. Analyses were adjusted for sex and the first ten genetic principal components, and p-values and confidence intervals were estimated using 10,000 bootstrapped simulations. Partial and complete mediation were declared following the definitions used in the discovery analysis.

## 3 Results

### 3.1 Characteristics of participants in the discovery GWAS

The discovery cohort comprised 12,806 participants of European ancestry and their characteristics are summarized in **Table 1**. Furthermore, we presented their characteristics stratified by case status for each NPS in **Supplementary Table 3**. The cohort had relatively similar representation of males and females (52% versus 48%, respectively). About 77% of participants experienced dementia during the observation period. Hallucinations and delusions had the lowest case rates, at under 25%, while the case rate was about 40% for disinhibition and over 60% for anxiety, apathy, depression/dysphoria, and irritability/lability. For all nine NPS domains, NPS symptom severity was positively correlated with severity of cognitive impairment measured by the global CDR score, with Spearman rho ranging from 0.08 to 0.50 (**Supplementary Table 4).** NPS symptom severity was negatively correlated with age of onset of cognitive impairment, with correlation coefficients ranging from -0.07 to -0.21 (**Supplementary Table 4**).

**Table 1.** Characteristics of the Alzheimer’s Disease Research Centers (ADRCs) participants in the discovery cohort (N=12,806).

|  | N | % |  |
| --- | --- | --- | --- |
| Cognitive status |  |  |  |
| MCI | 2,962 | 23% |  |
| dementia | 9,844 | 77% |  |
| Sex |  |  |  |
| female | 6,168 | 48% |  |
| male | 6,638 | 52% |  |
|  | N cases | % cases | N missing |
| Agitation or aggression | 7,015 | 55% | 78 |
| Anxiety | 7,894 | 62% | 79 |
| Apathy or indifference | 7,895 | 62% | 79 |
| Delusions | 3,081 | 24% | 83 |
| Depression or dysphoria | 7,581 | 60% | 79 |
| Disinhibition | 5,070 | 40% | 78 |
| Hallucinations | 2,027 | 16% | 81 |
| Irritability or lability | 7,863 | 62% | 78 |
| Nighttime behaviors | 7,085 | 56% | 90 |
|  | mean (SD) | median | range |
| Age at baseline visit | 73.1 (10.0) | 74 | 22.0–110.0 |
| Education (years) | 15.6 (2.9) | 16 | 0–30 |
| Number of visits | 4.6 (3.2) | 4 | 1–18 |

### 3.2 Genome-wide association analysis in each of the nine NPS

We performed a GWAS of each of the nine NPS, adjusting for sex, batch, and 10 genetic principal components. All GWAS had genomic inflation factor estimates of less than 1.02, suggesting no evidence of inflation (QQ plots in **Supplementary Figure 1**). We found GWAS-significant SNPs for agitation, anxiety, apathy, delusions, and hallucination. In all of these five NPS domains, the strongest signal overlaps the gene *APOE* (**Figure 1a**) and the lead SNP is rs429358 (Table 2). The C allele at this SNP is linked to the *APOE* ε4 allele and associated with increased risk for the five NPS. We tested whether the significant *APOE* SNP associations were modified by sex by performing a regression analysis adding a SNP-by-sex interaction term. We found SNP-by-sex interaction at unadjusted p-value < 0.05 for all traits except anxiety, but none of the interactions were significant after multiple testing correction for five traits, suggesting no detectable interaction between rs429358 and sex for these NPS symptoms. For agitation, a second significant locus comprising one SNP was observed on chromosome 9 in the gene *ADAMTS1L1* (**Figure 1b**, **Table 2**).

**Figure 1.**
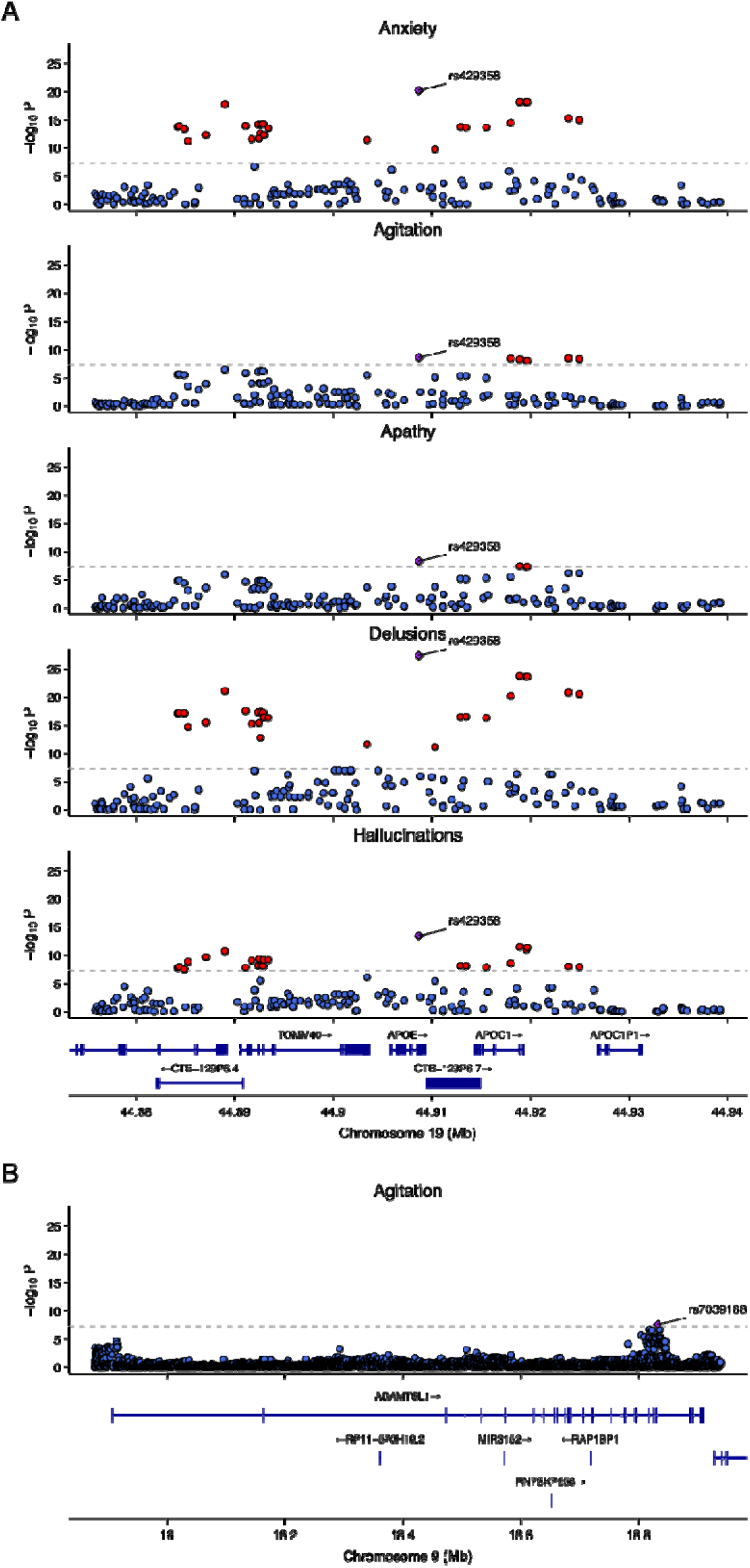
Regional P-value plots. Purple diamond indicates lead SNP in locus. Red points indicate SNPs passing the genome-wide significant (GWS) threshold of 5 × 10–8, and blue points indicate SNPs with p-values below the GWS threshold. A. Region on chromosome 19 with GWS SNPs n five NPS domains. B. Chromosome 9 region with GWS SNP in agitation.

**Table 2.** Lead SNPs at genomewide-significant loci.

ble 2. Lead SNPs at genomewide-significant loci.
| Locus | Lead SNP | Position | A1 | A2 | MAF | Closest Gene | Trait | OR (95% CI) | p-value |
| --- | --- | --- | --- | --- | --- | --- | --- | --- | --- |
| 1 | rs429358 | chr19:44,908,684 | C | T | 0.28 | APOE | Delusions | 1.42 (1.33, 1.51) | 4.5E-28 |
|  |  |  |  |  |  |  | Anxiety | 1.32 (1.24, 1.40) | 5.9E-21 |
|  |  |  |  |  |  |  | Hallucinations | 1.32 (1.23, 1.42) | 3.5E-14 |
|  |  |  |  |  |  |  | Agitation | 1.18 (1.12, 1.25) | 2.3E-09 |
|  |  |  |  |  |  |  | Apathy | 1.19 (1.12, 1.26) | 4.6E-09 |
| 2 | rs7039188 | chr9:18,833,016 | A | G | 0.26 | ADAMTSL1 | Agitation | 1.18 (1.11, 1.25) | 2.6E-08 |

A1, effect allele; A2, other allele; MAF, minor allele frequency in the dataset; OR, odds ratio; CI, confidence interval.

### 3.3 Mediation analysis of GWAS signals by severity and onset age of cognitive impairment

APOE variation, especially *APOE* ε4 allele count, is a known risk factor for Alzheimer’s disease. We tested whether the *APOE* GWAS signal for NPS was be mediated by cognitive impairment. We first quantified the proportion of the SNP effects that are mediated by age of onset of cognitive impairment. Since the significant SNPs in the APOE locus are in high LD with each other, we focused on the effect of lead SNP rs429358. We found that age of onset of cognitive impairment partially mediates the SNP effect on NPS, with mediation proportion of about 53% for apathy, 43% for agitation, and 13-35% for anxiety, hallucinations, and delusions (**Figure 2, Supplementary Table 5**). Next, we quantified the proportion of the SNP effects mediated by cognitive impairment severity. We found that cognitive impairment severity represented by binary MCI/dementia diagnosis completely mediates the SNP association with apathy but only partially mediates the SNP association with agitation, anxiety, hallucinations, and delusions (**Supplementary Table 5)**. Specifically, MCI/dementia diagnosis explains an estimated 61% of the SNP effect for agitation and 30-40% for anxiety, hallucinations, and delusions (**Figure 3A**). Results are similar when cognitive impairment severity is represented by global CDR score instead of MCI/dementia status, with partial mediation for all traits except apathy and agitation **(Figure 3B, Supplementary Table 5**). Overall, the mediation analysis suggests that for anxiety, hallucinations, delusions, and agitation the *APOE* locus signal is only partially mediated by the association of *APOE* ε4 with cognitive impairment.

**Figure 2.**
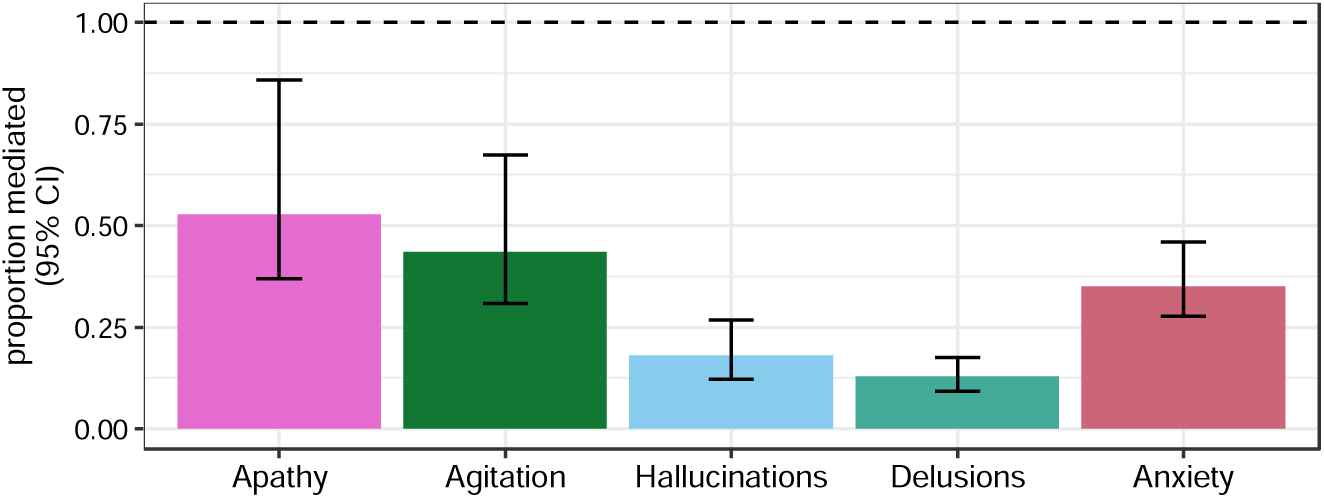
Proportion of APOE lead SNP effect mediated by age of onset of cognitive decline. The proportion mediated was estimated using R package mediation. Each bar corresponds to one of the five NPS with significant GWAS SNPs. Error bars indicate 95% confidence interval.

**Figure 3.**
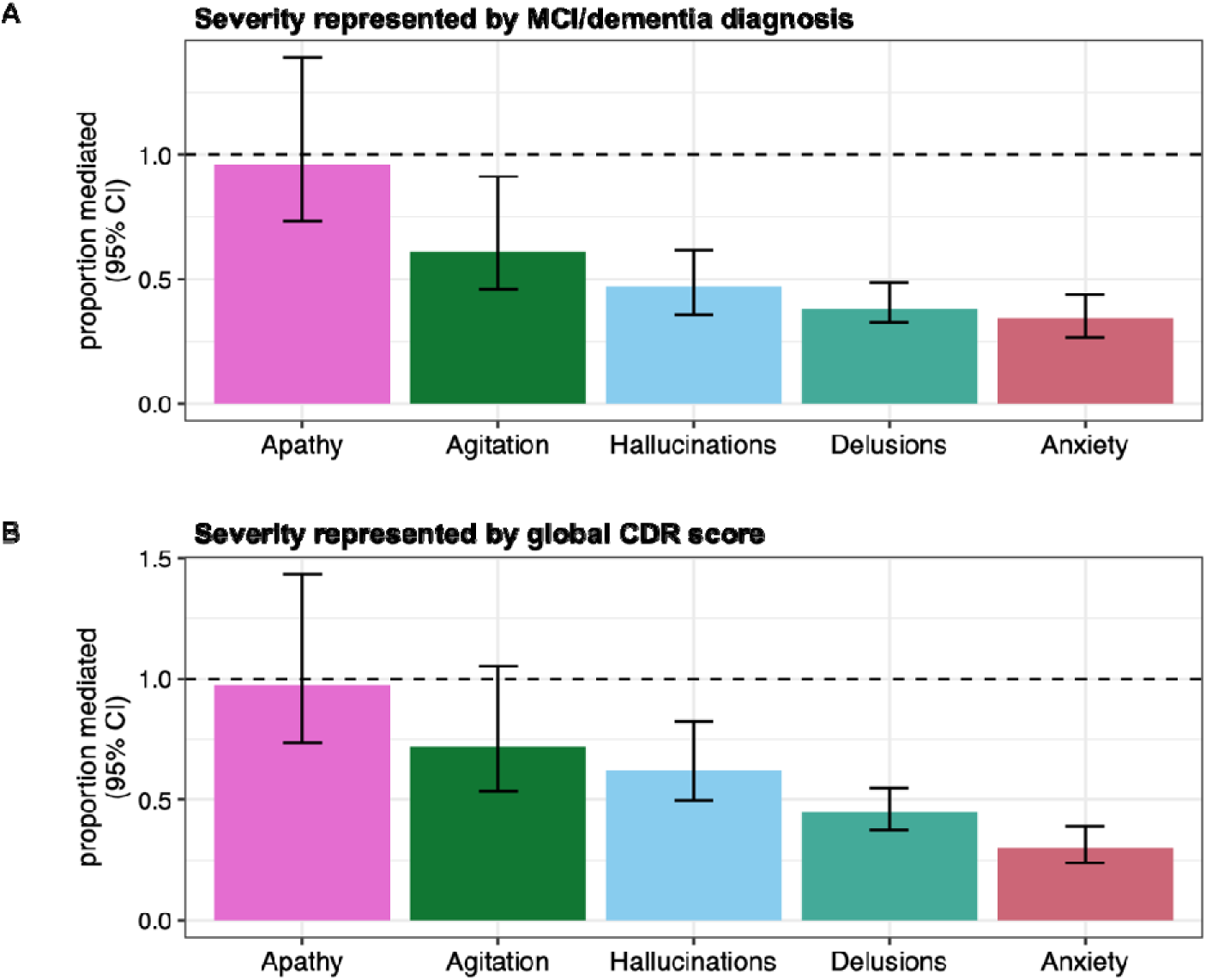
Proportion of APOE lead SNP effect mediated by cognitive impairment severity. The proportion mediated was estimated using R package mediation. Each bar corresponds to one of the five NPS with significant GWAS SNPs. Error bars indicate 95% confidence interval. A. Proportion mediated by cognitive impairment severity represented by binary MCI/dementia diagnosis. B. Proportion mediated by cognitive impairment severity represented by global CDR score.

### 3.4 Replication of GWAS findings in BioVU cohort

#### 3.4.1 Characteristics of VUMC BioVU replication cohort

We conducted a replication analysis using Vanderbilt University Medical Center’s (VUMC) genetic biobank, BioVU, and accompanying deidentified electronic health records. There were 2,456 BioVU participants of European genetic ancestry who had a first MCI/dementia ICD code occurring at or after age 50 and therefore met our criteria for inclusion. Sample characteristics are summarized in **Table 3**.

**Table 3.** Demographic and medical characteristics of European ancestry participants in a replication sample derived from the Vanderbilt University Medical Center genomic biobank (BioVU) (N=2,456)

|  | **Median (IQR)** |
| --- | --- |
| Birth year | 1940 (1933, 1947) |
| Sex |  |
| Female | 1,230 (50%) |
| Male | 1,226 (50%) |
| Medical record length (years) | 14 (8, 19) |
| Median age across medical record (years) | 74 (68, 81) |
|  | **N (%)** |
| Cognitive status (derived from diagnostic codes) |  |
| Dementia | 2,055 (84%) |
| Mild cognitive impairment (MCI) | 401 (16%) |
| Age at first MCI/dementia code | 75 (68, 81) |
| Antidepressant medication status |  |
| No antidepressant medication in record | 544 (22%) |
| First medication record > 1 year before first MCI/dementia ICD code | 1,273 (52%) |
| First medication record within 365 days or after MCI/dementia ICD code | 639 (26%) |
| Antipsychotic medication status |  |
| No antipsychotic medication in record | 1,255 (51%) |
| First medication record > 1 year before first MCI/dementia ICD | 404 (16%) |
| First medication record within 365 days or after MCI/dementia ICD code | 797 (32%) |
| Any neuropsychiatric symptom<sup>1</sup> |  |
| No antipsychotic or antidepressant medication | 350 (14%) |
| First medication record > 1 year before MCI/dementia ICD | 970 (39%) |
| First medication record within 365 days or after MCI/dementia ICD code | 1,136 (46%) |

<sup>1</sup>Binary variable based on first recorded receipt of medication within 365 days prior to or any time after first MCI/dementia ICD code Note: IQR: interquartile range, ICD: international classification of disease

To validate the use of ICD codes for selecting MCI/dementia participants in BioVU, we examined the association between APOE ε4 and ε2 isoforms (the strongest genetic risk and protective factors, respectively, for AD dementia) and BioVU MCI/dementia status. We performed propensity score matching on median age across record and sex to obtain the controls for the MCI/dementia cases. Association analysis showed that the MCI/dementia participants were more likely to carry the ε4 allele than matched controls (38% versus 26% ε4 carriers, p-value<0.001) and less likely to carry the ε2 allele (13.5% versus 15.6%, p-value=0.04), validating the selection of the MCI/dementia cases. Of the 2,456 MCI/dementia participants, 1,136 were cases for any NPS based on receipt of antipsychotic and/or antidepressant medications (**Supplementary Table 6**).

#### 3.4.2 Replication of SNP association

In the discovery GWAS, one SNP in *ADAMTSL1* and 27 SNPs in or near *APOE* met the genome-wide significance threshold for at least one NPS (before LD pruning). The *ADAMTSL1* SNP and 24 of the *APOE* SNPs were available in the BioVU replication cohort. Since the SNPs in the *APOE* locus are in high LD, for purposes of defining the number of independent tests, we considered these as essentially the same signal and used Bonferroni multiple testing correction for the two independent tested loci, setting the significance threshold at 0.05 / [2 independent loci] = 0.025.

In the *APOE* locus,14 of 24 SNPs replicated (**Supplementary Table 7**), including the lead SNP from the discovery analysis (**Figure 4**). In the *ADAMTSL1* locus, the significant SNP from the discovery GWAS was significantly associated with the any NPS outcome (p-value = 7.8×10^-5^, **Supplementary Table 7**), but the direction of association was opposite to that observed in the discovery GWAS (**Figure 4**).

**Figure 4:**
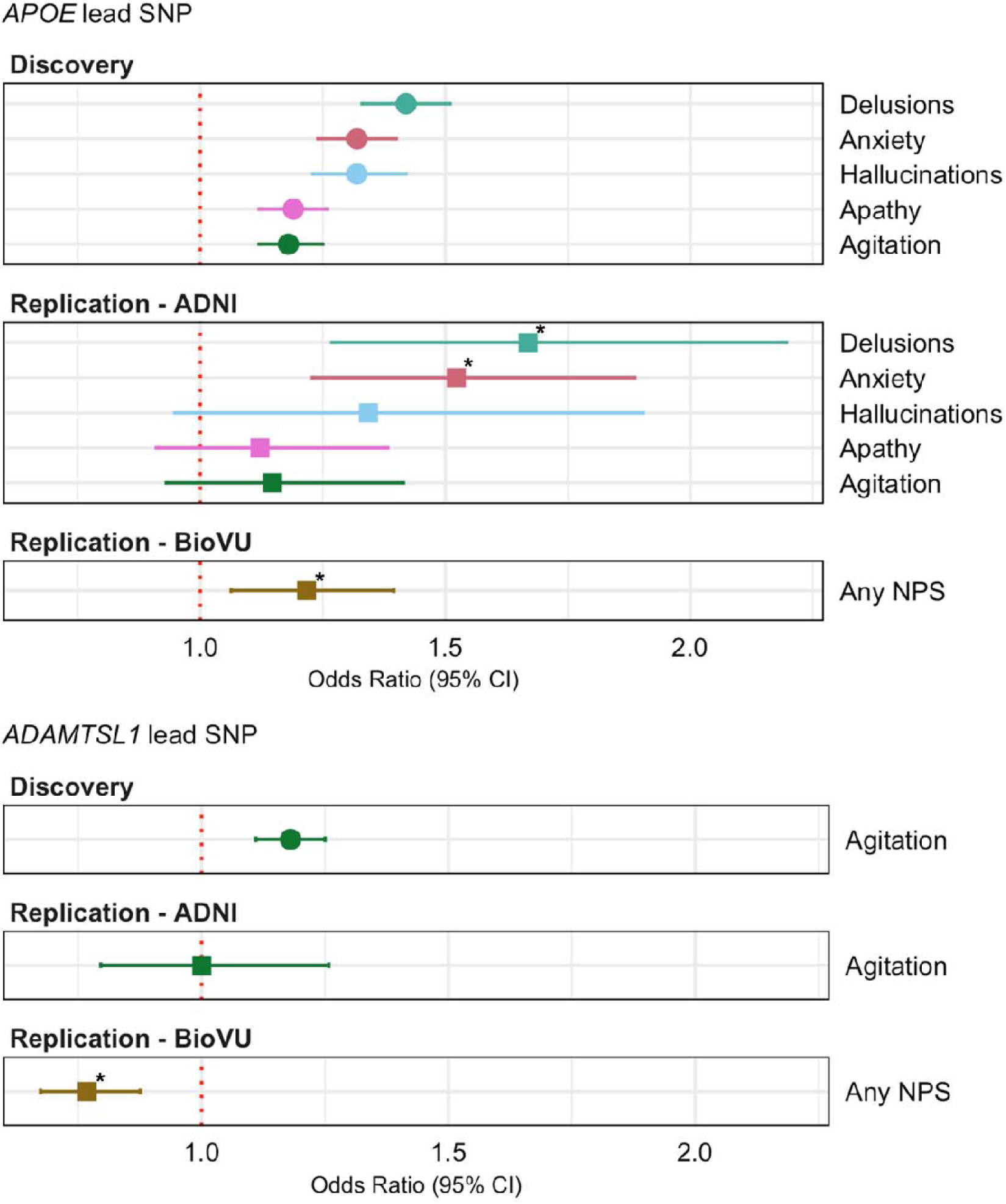
Odds ratio estimates for lead SNPs in discovery and replication cohorts. Odds ratio estimates are from the NACC discovery analysis (filled circles) or BioVU or ADNI replication analyses (filled squares). The NPS outcomes in the discovery a alysis and ADNI analysis were defined using NPI or NPI-Q responses. The any NPS outcome in the BioVU analysis was defined using receipt of antipsychotic and/or antidepressant medication within one year before or any time after a patient’s first MCI or dementia ICD code. Asterisks for replication results indicate effects that are significant at alpha = 0.05 after Bonferroni correction for the number of independent tests performed in each analysis (two tests in BioVU, six tests in ADNI).

#### 3.4.3 Mediation analysis

Since the *APOE* SNP associations largely replicated in BioVU, we conducted mediation analyses to examine whether these associations are mediated by MCI/dementia. For mediators, we considered (i) the age at first MCI/dementia diagnosis and (ii) MCI vs. dementia status. We found no mediation by age at first MCI/dementia diagnosis (**Supplementary Table 8**). The binary MCI versus dementia diagnosis variable partially mediated the association between the SNPs and NPS, with the proportion mediated ranging from 12% to 22% (**Supplementary Table 8**).

### 3.5 Replication of GWAS findings in ADNI cohort

#### 3.5.1 Characteristics of the ADNI cohort

We conducted a second replication analysis in the ADNI cohort. After filtering to those of European ancestry with MCI or dementia, there were 798 participants available for analysis. About 41% of the participants were female, and almost half exhibited dementia during the observation. Additional sample characteristics are summarized in **Table 4**, and similar tables stratified by NPS status are presented in **Supplementary Table 9**.

**Table 4.**
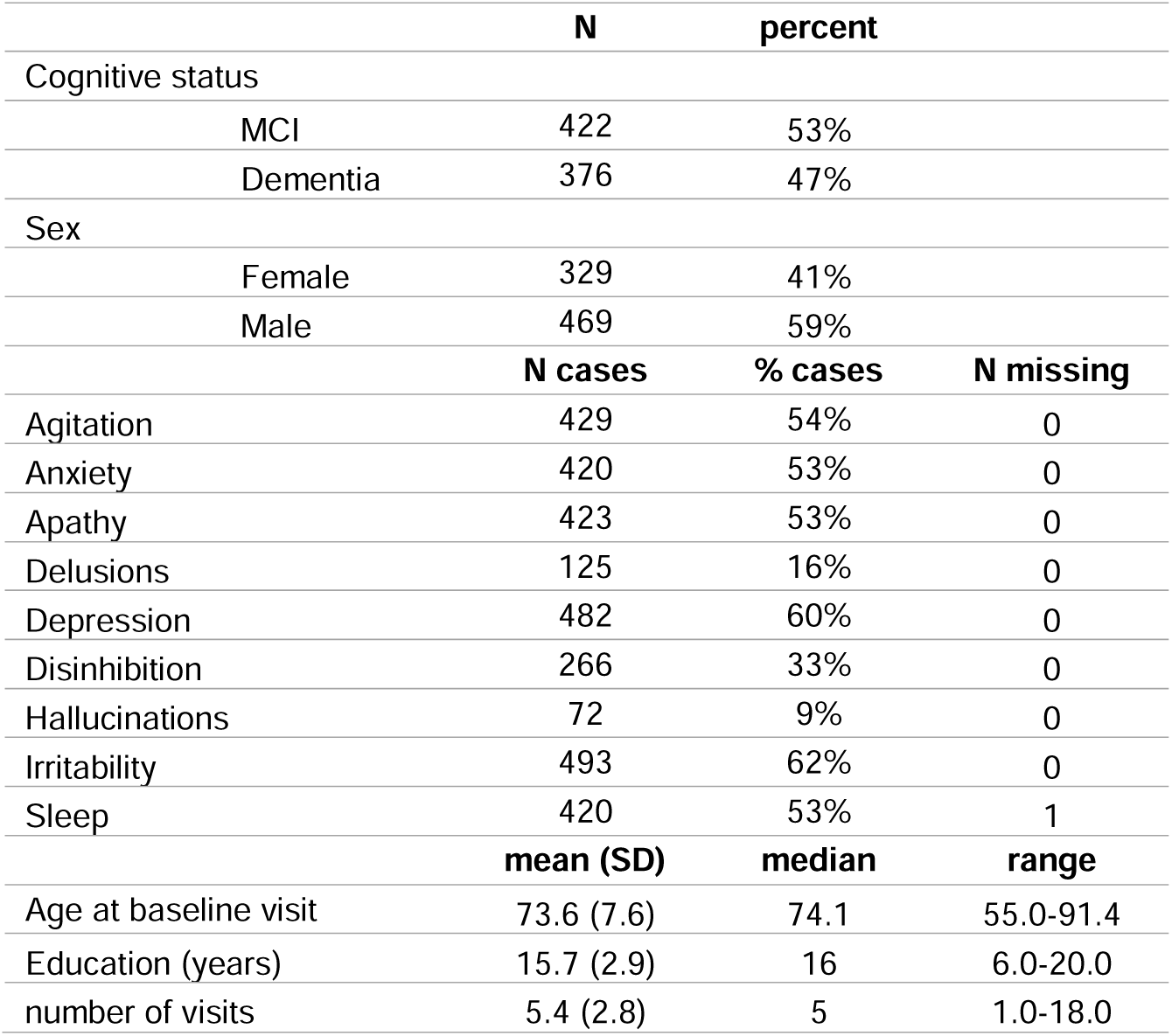
Characteristics of ADNI replication cohort (N=798).

#### 3.5.2 Replication of SNP association

Of the 28 SNPs from the *ADAMTSL1* and *APOE* loci that met the genome-wide significance in the discovery GWAS (before LD pruning), the *ADAMTSL1* SNP and 24 of the *APOE* SNPs were available in the ADNI replication cohort. Like our rationale in the BioVU replication, we considered the *APOE* SNPs to represent a single locus and used Bonferroni correction for six independent tests (the *ADAMTSL1* test for agitation and five NPS tested for *APOE*), setting the significance threshold at 0.05/ [6 tests] = 8.33 × 10^-3^.

In the *APOE* locus, all 24 of the SNPs tested for anxiety and 18 of the 24 SNPs tested for delusions were significantly associated after Bonferroni correction (**Supplementary Table 10**). The SNP rs429358 was significantly associated with both traits (**Figure 5**). While some *APOE* SNPs had nominally significant association with hallucinations, they did not survive Bonferroni correction. No association was observed for agitation or apathy, possibly due to a combination of low statistical power and smaller effect sizes. Likewise, the *ADAMTSL1* SNP associated with agitation in the discovery analysis was not significantly associated with agitation in the ADNI replication test.

#### 3.5.3 Mediation analysis

We performed mediation analysis for the two outcomes with significant SNP association for comparison with the mediation results in the discovery analysis. We found that severity of cognitive impairment represented by global CDR score or binary MCI/dementia status only partially mediated the SNP effect on anxiety and delusions, consistent with the findings in the discovery cohort.

Specifically, the estimated proportion of the SNP effect mediated by global CDR score was 39% (95% CI [0.20, 1.00]) for delusions and 27% (95% CI 0.11, 0.67]) for anxiety. The confidence intervals were larger than the discovery analysis, which is expected due to the smaller sample size. We found no mediation by onset age of cognitive impairment in the association between *APOE* SNPs and anxiety or delusion in ADNI dataset (**Supplementary Table 11**). Together, the association between *APOE* SNPs and delusion and anxiety, respectively, was replicated in the ADNI dataset and these associations were beyond the effect of APOE SNPs on MCI / dementia severity.

## 4 Discussion

We conducted parallel GWAS of nine NPS in over 12,000 participants with mild cognitive impairment or ADRD, recruited by U.S. ADRCs. We identified two genome-wide significant loci for NPS, providing insights into the genetic underpinnings of these symptoms. The first locus, within the *ADAMTSL1* gene, was associated with agitation. The second and stronger locus, overlapping *APOE* and driven by the *APOE* ε4 allele, was associated with multiple NPS domains, including agitation, anxiety, apathy, delusions, and hallucinations. While *APOE* ε4 is well established as a risk factor for AD dementia, our mediation analysis suggests its association with most of the NPS domains is beyond its effects on AD dementia. The results therefore underscore that *APOE* ε4 influences NPS and cognitive impairment through partially distinct mechanisms. The associations with anxiety, delusions, and hallucination were the strongest and showed the least mediation by cognitive impairment severity, pointing to a more direct role of *APOE* in these symptoms.

We followed up on the findings from the discovery analysis using two independent replication datasets: the ADNI dataset of ∼800 participants where NPI/NPI-Q records characterized the same nine NPS domains, and the BioVU dataset of ∼2500 participants where a general NPS outcome was defined based on receipt of NPS-related medications. The *ADAMTSL1* locus replicated in BioVU but with reversed effect direction and did not replicate in ADNI, indicating that further evaluation in other datasets is needed to clarify the relationship between *ADAMTSL1* and agitation. In contrast, the *APOE* locus demonstrated consistent replication. The associations with anxiety and delusions were replicated in ADNI, while the general NPS outcome replicated in BioVU. Despite challenges in sample size and phenotype matching, our replication analyses in ADNI and BioVU provide support for the *APOE* associations observed in the discovery analysis, reinforcing the robustness of these findings.

Prior studies have examined the effect of *APOE* variation on NPS risk in cognitively impaired individuals, although in many cases the studies have had modest sample sizes. For instance, a recent meta-analysis^22^ reported a median sample size of just 194 across 53 studies, with 75% of studies including fewer than 400 participants. Variations in NPS definitions, sample characteristics (e.g., age, dementia status), and methodology also present challenges. Nevertheless, they present valuable preliminary evidence, and we summarize the existing literature in the next paragraphs.

A 2011 study of ∼2300 people with dementia^42^ found no association between *APOE* ε4 and psychosis. Similarly, a 2021 GWAS meta-analysis of ∼12.3K people with Alzheimer’s disease^25^ found no genome-wide significant *APOE* variants for psychosis; however, gene-based analysis of the same data found a positive association signal linked to the ε4 allele^25^, as did an older candidate gene study in 501 individuals with AD^43^. Our observed association of *APOE* genetic variation with delusions and hallucination aligns with these prior findings.

For apathy, a study of 201 people with AD found that *APOE* ε4 carriers had increased risk for syndromes that included apathy^44^. However, the relationship among *APOE*, cognitive impairment, and apathy is uncertain. Interestingly, in *APOE* ε4 carriers without dementia, apathy was associated with a lower risk of incident dementia, possibly because *APOE* ε4 confers a strong baseline risk for dementia that is proportionally large compared to any independent association of apathy with dementia^45^. Consistent with this, we found that *APOE* ε4 was positively associated with apathy, but the effect was fully mediated by cognitive impairment severity. Our results complement existing findings and suggest that apathy is a consequence of cognitive decline rather than an independent symptom.

For agitation, studies of the association with *APOE* are limited. However, in individuals with MCI, agitation was found to strongly interact with *APOE* ε4 carrier status and increase the risk for incident dementia^46^. This study also found *APOE* interacts with anxiety, apathy, irritability, depression, and nighttime behaviors on incident dementia risk. Our observations are consistent in suggesting *APOE* ε4 is linked to agitation and other NPS by mechanisms that are not directly derived from cognitive impairment.

Unlike previous candidate gene studies, our work employed a genome-wide approach, allowing for agnostic discovery of *APOE* association with NPS. Some existing GWAS studies have investigated subsets of NPS domains. A 2021 GWAS meta-analysis of psychosis^25^ in AD found two genome-wide significant SNP association signals, in *ENPP6* and *SUMF1*. A 2024 GWAS meta-analysis^24^ in ∼10,000 participants by the same group found no genomewide-significant loci, but highlighted a locus near *RAD23B* that approached significance (lead SNP p-value p-value=1.33×10^−7^) for affective disturbance. None of these loci were significant in our analysis, but the lack of consistency can be explained by multiple factors. For instance, we did not consider precisely the same NPS outcome.

*APOE* is well established as a key player in neuroinflammation. Emerging evidence from *in vivo* PET imaging of TSPO protein (a marker of microglial activation) shows that neuroinflammation in the human brain is associated with neuropsychiatric symptoms and, importantly, the associations persist after accounting for confounders like amyloid and tau burden and cognitive impairment severity^47,48^. These observations align with previous work showing that inflammation contributes to risk and severity of psychiatric conditions such as PTSD^49^ and depression^50^. These insights together with our observations strengthen the hypothesis that *APOE* ε4 influences NPS through its role in neuroinflammation, opening avenues for future research into inflammation-targeted therapies for managing NPS in dementia.

Our study has some limitations. First, while our study represents one of the largest GWAS of NPS to date, even larger sample sizes will be essential to detect genetic variants with small effect sizes and uncover additional loci. Second, the complex dependencies among *APOE* variation, cognitive impairment, and NPS mean that our result may be specific to the population, setting and model formulation used. For instance, NACC participants tend to be highly educated, and our analysis focused on only individuals of European ancestry. Third, the BioVU replication analysis relied on a medication-based NPS outcome, which lacks the domain-specificity of the NPI-Q outcomes used in the discovery analysis. Fourth, the ADNI replication dataset had small sample size, and therefore may not have been well-powered to replicate true but modest associations.

Future work could explore alternate case definitions and leverage multivariate analysis or factor analysis to better account for substructure in the NPS domains. Sex-stratified analyses may also identify genetic variants with sex-specific effects, especially if larger cohorts can be assembled to maintain power.

In summary, our findings underscore the multifaceted role of *APOE* ε4, influencing both cognitive impairment and NPS likely through partially distinct mechanisms, and suggest future research should investigate APOE-related mechanisms and therapeutic targets to better understand and manage neuropsychiatric symptoms in MCI and ADRD.

## 5 Data Availability

All datasets used in this study are under controlled access to protect patient privacy. ADGC genotype data may be requested from the Alzheimer’s Disease Genetics Consortium (https://www.adgenetics.org) by submitting a Special Analysis Group (SAG) proposal. UDS phenotypes may be requested from NACC (https://naccdata.org) by submitting a data request. Access to the BioVU resource is available to Vanderbilt faculty members and their collaborators who have an IRB approved project, sign a data use agreement, and receive permission of protocol from the Vanderbilt Institute for Clinical and Translation Research (VICTR-BioVU) review committee. Access to ADNI data can be requested by accepting the data use agreement and submitting a data use application to the ADNI Data Sharing and Publications Committee. Summary statistics generated in this study are available from NIAGADS.

## 6 Code Availability

The analysis was performed using publicly available software as described in Methods: vcftools (0.1.13); bcftools (1.19); tabix; KING (2.3.0); Eigenstrat (6.1.4); PLINK 1.9; PLINK2; R (3.6 and 4.4); R package mediation (4.5.0). Code is available on request.

## Data Availability

All data produced in the present study are available upon reasonable request to the authors.

## 7 Acknowledgements

This work was supported by the following grants to the authors: R01 AG075827, R01 AG072120, IK4BX005219 to APW and TSW; R01 AG079170 to TSW; T32 HG008341 to FB; 1R01MH118233 to LKD. TWM receives funding from the Tourette Association of America. The Alzheimer’s Disease Genetics Consortium supported the collection of samples used in this study through National Institute on Aging (NIA) grants U01AG032984 and RC2AG036528. The NACC database is funded by NIA/NIH Grant U24 AG072122. VUMC BioVU is supported by numerous sources including the NIH funded Shared Instrumentation Grant S10RR025141 and CTSA grants UL1TR002243, UL1TR000445, and UL1RR024975. Data collection and sharing for this project was funded by the Alzheimer’s Disease Neuroimaging Initiative (ADNI) (National Institutes of Health Grant U01 AG024904) and DOD ADNI (Department of Defense award number W81XWH-12-2-0012). ADNI is funded by the National Institute on Aging, the National Institute of Biomedical Imaging and Bioengineering, and through generous contributions from the following: AbbVie, Alzheimer’s Association; Alzheimer’s Drug Discovery Foundation; Araclon Biotech; BioClinica, Inc.; Biogen; Bristol-Myers Squibb Company; CereSpir, Inc.; Cogstate; Eisai Inc.; Elan Pharmaceuticals, Inc.; Eli Lilly and Company; EuroImmun; F. Hoffmann-La Roche Ltd and its affiliated company Genentech, Inc.; Fujirebio; GE Healthcare; IXICO Ltd.; Janssen Alzheimer Immunotherapy Research & Development, LLC.; Johnson & Johnson Pharmaceutical Research & Development LLC.; Lumosity; Lundbeck; Merck & Co., Inc.; Meso Scale Diagnostics, LLC.; NeuroRx Research; Neurotrack Technologies; Novartis Pharmaceuticals Corporation; Pfizer Inc.; Piramal Imaging; Servier; Takeda Pharmaceutical Company; and Transition Therapeutics. The Canadian Institutes of Health Research is providing funds to support ADNI clinical sites in Canada. Private sector contributions are facilitated by the Foundation for the National Institutes of Health (www.fnih.org). The grantee organization is the Northern California Institute for Research and Education, and the study is coordinated by the Alzheimer’s Therapeutic Research Institute at the University of Southern California. ADNI data are disseminated by the Laboratory for Neuro Imaging at the University of Southern California.

## 8 Author Contributions

APW and TSW conceived the study. SMV performed analysis in the discovery and ADNI datasets, and FB performed analysis in the BioVU dataset. APW, TSW, TMF, SMV, and FB interpreted the results. SMV, FB, and APW wrote the initial manuscript. All authors revised and provided critical feedback on the manuscript. All authors approved the manuscript.

