## Supplementary figures and images for "GWAS links *APOE* to neuropsychiatric symptoms in mild cognitive impairment and dementia"

### Supplementary Figures

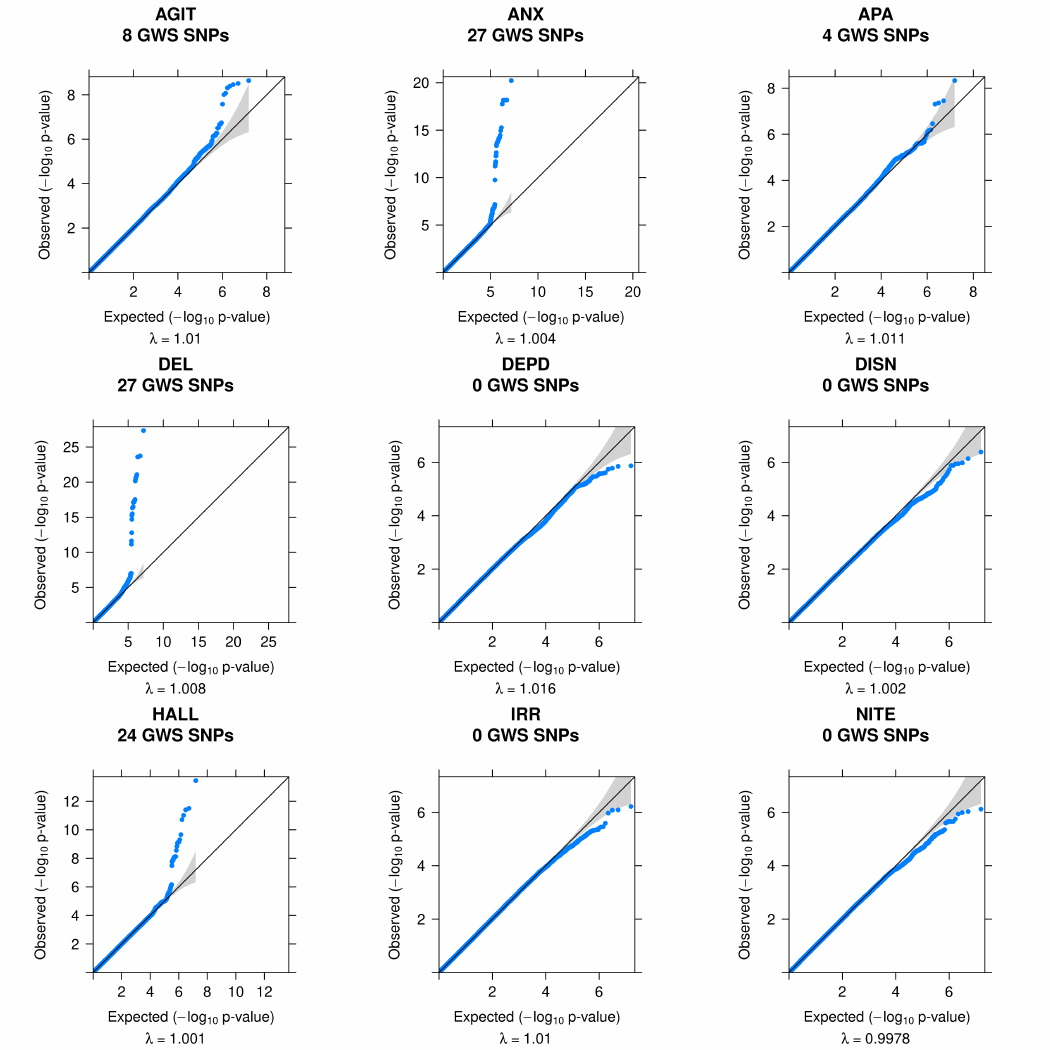

#### Supplementary Figure 1. QQ plots and lambda estimates from GWAS p-values per NPS.
